# Genetic Risk for Attention-Deficit/Hyperactivity Disorder Predicts Cognitive Decline and Development of Alzheimer’s Disease Pathophysiology in Cognitively Unimpaired Older Adults

**DOI:** 10.1101/2022.04.05.22273464

**Authors:** Douglas T. Leffa, João Pedro Ferrari-Souza, Bruna Bellaver, Cécile Tissot, Pamela C. L. Ferreira, Wagner S. Brum, Arthur Caye, Jodie Lord, Petroula Proitsi, Thais Martins-Silva, Luciana Tovo-Rodrigues, Dana L. Tudorascu, Victor L. Villemagne, Annie Cohen, Oscar L. Lopez, William E. Klunk, Thomas K. Karikari, Pedro Rosa-Neto, Eduardo Zimmer, Brooke S.G. Molina, Luis Augusto Rohde, Tharick A. Pascoal, the Alzheimer’s Disease Neuroimaging Initiative

## Abstract

**Background:** Attention-Deficit/Hyperactivity Disorder (ADHD) persists in older age and is postulated to be a risk factor for cognitive impairment and Alzheimer’s Disease (AD). However, this notion relies exclusively on epidemiological associations, and no previous study has linked ADHD with a decline in cognitive performance in older adults or with AD progression. Therefore, this study aimed to determine whether genetic liability for ADHD, as measured by a well-validated ADHD polygenic risk score (ADHD-PRS), is associated with longitudinal cognitive decline and the development of AD pathophysiology in cognitively unimpaired (CU) older adults.

**Methods:** We calculated a weighted ADHD-PRS in 212 CU individuals without a clinical diagnosis of ADHD (55-90 years) using whole-genome information. These individuals had baseline amyloid-β (Aβ) positron emission tomography, as well as longitudinal cerebrospinal fluid (CSF) phosphorylated tau at threonine 181, structural magnetic resonance imaging, and cognitive assessments for up to 6 years. Linear mixed-effects models were used to test the association of ADHD-PRS with cognition and AD biomarkers.

**Outcomes:** Higher ADHD-PRS was associated with greater cognitive decline over 6 years. The combined effect between high ADHD-PRS and brain Aβ deposition on cognitive deterioration was more significant than each individually. Additionally, higher ADHD-PRS was associated with increased CSF p-tau_181_ levels and frontoparietal atrophy in CU Aβ-positive individuals.

**Interpretation:** Our results suggest that genetic liability for ADHD is associated with cognitive deterioration and the development of AD pathophysiology in the CU elderly. These findings indicate that ADHD-PRS might inform the risk of developing cognitive decline in this population.

**Funding:** National Institute of Health and Brain & Behavioral Research Foundation.

## Introduction

Attention-deficit/hyperactivity disorder (ADHD) is characterized by impairing and pervasive symptoms of inattention, hyperactivity-impulsivity, or both (1). Although first conceptualized as a neurodevelopmental disorder of childhood, current evidence supports ADHD as a waxing and waning condition with fluctuating symptoms and impairments throughout the lifespan (2), including in older adulthood (3). According to a recent meta-analysis, the prevalence of ADHD in older adults (>50 years) is approximately 2.18% (4). Furthermore, as the geriatric population grows, the absolute number of patients aged 50 years and older fulfilling the criteria for diagnosis of ADHD will likely increase (3). Therefore, understanding the disorder’s association with prevalent age-related diseases is a pressing concern.

The association between ADHD and age-related cognitive impairment is of particular interest. Throughout the lifespan, cognitive deficits across various neurocognitive domains have been extensively described in ADHD (1). Additionally, cognitive function in older adults with ADHD may closely resemble early manifestations of neurodegenerative conditions (5). Recent population-based large epidemiological studies suggested that ADHD is associated with a higher risk for mild cognitive impairment (MCI) and Alzheimer’s disease (AD) (6–8). However, these findings rely primarily on electronic health records and consequently can present biased estimates of ADHD and dementia prevalence. Since the differential diagnosis between undiagnosed ADHD and early dementia can be challenging (5), it is crucial to clarify whether ADHD is a risk factor for MCI and AD dementia or misdiagnosis due to symptom overlap (5). Additionally, it is unclear whether ADHD is associated with progressive cognitive decline in older age, above and beyond the cognitive deficits originating in childhood (1).

An obstacle to investigating age-related cognitive decline in ADHD is the absence of large-scale studies following patients with childhood-diagnosed ADHD into older age. An alternative approach is to consider the association of well-established dimensional biomarkers of ADHD and AD in samples not selected for ADHD. The ADHD polygenic risk score (ADHD-PRS) represents the combined genetic liability for the disorder and is highly associated with ADHD diagnosis and related traits in independent clinical and population samples (9). In this study, we explored the association of ADHD-PRS with cognitive impairment in older age. More specifically, we tested the following hypotheses: (1) ADHD-PRS is associated with progressive cognitive decline in cognitively unimpaired (CU) older people; (2) the association between ADHD-PRS and cognitive decline varies according to baseline Aβ burden; (3) ADHD-PRS is associated with brain tau pathology and degeneration.

## Methods

### Participants

We used data from the Alzheimer’s Disease Neuroimaging Initiative (ADNI), a longitudinal multicenter study designed to develop clinical, imaging, genetic, and biochemical biomarkers for the early detection and tracking of AD (http://adni.loni.usc.edu; for more information, see previous reports (10)). ADNI’s inclusion criteria relevant for this study are age between 55 and 90, absence of major depression or bipolar disorder (DSM-IV criteria) within the past one year, no history of schizophrenia (DSM-IV criteria), and a Geriatric Depression Scale (GDS) score less than 6. All data were downloaded from the ADNI data repository in December 2021. Institutional Review Boards of all involved sites approved the ADNI study, and all research participants or their authorized representatives provided written informed consent.

We included CU participants with baseline medical data, baseline Aβ [^18^F]florbetapir positron emission tomography (PET), whole-genome information, and a minimum of two clinical assessments with neuropsychological testing. At baseline, all CU participants included in the analysis had a Clinical Dementia Rating (CDR) score of 0 and a Mini-Mental State Exam (MMSE) score of 24 to 30. After the baseline assessment, subjects returned at six months, one year, and annually after that (mean number of observations with complete data = 3.6, with a minimum of 2 and a maximum of 7 observations per individual). Analyses evaluating the longitudinal trajectory of (CSF) tau phosphorylated at threonine 181 (p-tau_181_) and gray matter (GM) volume were performed in a subsample based on data availability (Figure S1).

### Polygenic risk score

Data were available across three genotyping platforms: (1) the Human610-Quad platform, (2) the HumanOmniExpress, and (3) Omni 2.5M platform. Imputation and merging of the different platforms were performed as previously described (11). ADHD-PRS was calculated using the additive model, which is the weighted sum of risk alleles for ADHD according to the most recent genome-wide association study (GWAS) (12). The calculation was performed using the PRSice software v2.2 (13). Independent single nucleotide polymorphisms (SNPs) were classified based on a 250-kb window and 0.1 r2 linkage disequilibrium criteria. After applying the quality control filters, 212,846 variants were retained for PRS analysis. Only genes located outside the MHC region (chr6: 26-33Mb) were included. Nine ADHD-PRSs were calculated using subsets of SNPs selected according to the following GWAS p-value thresholds: 1, 0.5, 0.4, 0.3, 0.2, 0.1, 0.05, 0.005, and 0.0005. Main analyses were performed with the threshold of 1, assuming all genetic markers contributed to ADHD diagnosis. ADHD-PRSs were transformed into z-scores for better visualization. To investigate populational structure, principal components analysis was conducted using PLINK 1.9 (14). We retained seven principal components to account for any ancestry differences in genetic structure that could bias the results, as previously done for ADNI datasets (11).

### Cognitive function

Each participant from ADNI was submitted to a broad clinical and neuropsychological assessment in selected visits. Our primary outcome was cognitive function as measured by the Preclinical Alzheimer’s Cognitive Composite (PACC) adapted in the ADNI study (15). The PACC was developed to detect the first signs of cognitive decline in CU subjects with biomarker evidence of AD pathology. The PACC adapted for ADNI was obtained by summing the following four standardized z-scores: Alzheimer Disease Assessment Scale - Cognitive Subscale Delayed Word Recall, Logical Memory Delayed Recall, MMSE, and Trail-Making Test B Time to Completion (15). Higher scores in PACC indicate better cognitive performance. To explore specific aspects of cognitive function, we used the ADNI composite score for executive function (ADNI-EF) (16) and the ADNI composite score for memory function (ADNI-Mem) (17). The ADNI-EF includes the performance on WAIS-R Digit Symbol Substitution, Digit Span Backwards (Trails A and B), Category Fluency, and Clock Drawing (16). The ADNI-Mem includes the Rey Auditory Verbal Learning Test, AD Assessment Schedule – Cognition, MMSE, and Logical Memory (17). For both, higher scores indicate better performance.

### MRI and PET

Magnetic resonance imaging (MRI) and [^18^F]florbetapir Aβ-PET were acquired following ADNI protocols and pre-processed as previously described (18). Briefly, MRIs were segmented into probabilistic GM maps using the SPM12 segmentation tool. Each GM probability map was then non-linearly registered (with modulation) to a stereotaxic space using DARTEL and smoothed with a Gaussian kernel of full-width half maximum of 8 mm to generate GM density voxel-based morphometry (VBM) images. We visually inspected all images to ensure proper alignment to the ADNI template. [^18^F]Florbetapir standardized uptake value ratio (SUVR) images used the whole cerebellum as the reference region and were generated from a weighted average of the mean uptake from the cortical GM of frontal, anterior and posterior cingulate, lateral parietal, and temporal regions (19). Individuals with [^18^F]florbetapir SUVR higher than 1.11 were classified as Aβ-positive, a widely validated cutoff for this population (19).

### CSF p-tau_181_

CSF p-tau_181_ was measured using fully automated Elecsys immunoassays (Roche Diagnostics). Measurements outside the analytical range (< 8 pg/mL or > 120 pg/mL) were handled by setting them to the lower or upper detection limit. One individual presenting CSF p-tau_181_ concentrations three standard deviations above the mean was considered an outlier and excluded from the analyses.

### Statistical analysis

Statistical analyses were performed using Stata version 14.0 (StataCorp, College Station, Texas USA) and voxel-wise statistics using MATLAB software version 9.2 (http://www.mathworks.com) with VoxelStats package (20). We used random field theory (RFT) to correct brain imaging results for multiple comparisons. The association between ADHD-PRS and baseline demographic characteristics was explored using linear or logistic regressions. A two-sided p-value lower than 0.05 was considered statistically significant.

Linear mixed-effects models were used to assess the main effects of ADHD-PRS on cognitive function (Model 1, Supplementary Material). To test the hypothesis that ADHD-PRS is associated with progressive cognitive decline, we examined the interaction between ADHD-PRS and time (Model 2, Supplementary Material). To test whether the association between ADHD-PRS and cognitive decline varies according to baseline Aβ burden, we added an interaction between ADHD-PRS, time, and Aβ-PET (Model 3, Supplementary Material). Cohen’s *f*^2^ allows the estimation of the effect size within the context of mixed-effects linear models and was obtained as previously described (21). Cohen’s *f*^2^ ≥ .02, *f*^2^ ≥ .15, and *f*^2^ ≥ .35 represent small, medium, and large effect sizes, respectively (21).

In exploratory analyses, we tested the association between ADHD-PRS and longitudinal changes in CSF p-tau_181_ or VBM. For that, we evaluated the interaction between ADHD-PRS and time, also with linear mixed-effects models (Models 4 and 5, respectively, Supplementary Material). Following a significant triple interaction between ADHD-PRS, time, and Aβ-PET, analyses with CSF p-tau_181_ and brain atrophy were conducted stratifying individuals according to their Aβ status. All models were adjusted for sex, age at baseline, apolipoprotein E ε4 (*APOE* ε4) carriership status (carriers vs. non-carriers), years of education, and ancestry (using the first seven principal components, as previously performed for ADNI datasets (11)). Furthermore, as recommended for proper adjustment in genetic studies, we adjusted for the interaction between the independent variables and each covariate (22). Time was defined as years from baseline for each participant. Mixed models were fit including subject-specific random slopes and intercepts to cluster the multiple assessments per individual.

We conducted the primary analyses including all SNPs (ADHD-PRSs with a threshold of 1). Sensitivity analyses with additional thresholds were performed and can be found in the Supplementary Material. Additionally, we performed sensitivity analyses to explore the role of potential confounders previously associated with ADHD and AD dementia, such as vascular risk factors (VRFs) (1,23) and depression (1,23). VRF burden was assessed using a composite score, and a score equal to or higher than two was defined as elevated (24) (Supplementary Material). Depression symptoms were assessed using baseline GDS scores.

## Results

A total of 212 participants had genetic data and Aβ-PET measures at baseline (Figure S1). From those, 196 and 193 participants had CSF p-tau_181_ and MRI data, respectively (Figure S1). The mean (SD) age of the sample was 73.1 (5.96) years, 116 (54.7%) were women, and all self-reported being White (210 as Not Hispanic/Latino). The mean and median observation time was 3.96 and 4.05 years, respectively (SD = 1.5, interquartile range = 3.01 – 5.31, maximum follow-up of 6 years). Table 1 shows participants’ characteristics at baseline. ADHD-PRSs were normally distributed (Figure S2) and were not associated with age, sex, or *APOE* ε4 carrier status (Table S1). Higher ADHD-PRS was associated with decreased years of education (Table S1). Additionally, ADHD-PRS was not associated with length of follow-up, suggesting no attrition bias (Table S1).

**Table 1.** Baseline demographic and clinical characteristics of the population.

| | Overall<br>(N = 212) | Aβ-negative<br>(N = 137) | Aβ-positive<br>(N = 75) |
| --- | --- | --- | --- |
| Age, y, mean (SD) | 73.1 (5.9) | 72.2 (5.7) | 74.8 (5.9) |
| Sex, No. (%) |  |  |  |
| Female | 116 (54.7) | 64 (46.7) | 52 (69.3) |
| Male | 96 (45.3) | 73 (53.3) | 23 (30.7) |
| Race, No. (%) |  |  |  |
| White | 212 (100) | 137 (100) | 75 (100) |
| Ethnicity, No. (%) |  |  |  |
| Not Hispanic/Latino | 210 (99.1) | 136 (99.3) | 74 (98.7) |
| Unknown | 2 (0.9) | 1 (0.7) | 1 (1.3) |
| Years of education, mean (SD) | 16.6 (2.5) | 16.8 (2.4) | 16.2 (2.6) |
| APOE ε4, No. carriers (%) | 63 (29.7) | 28 (20.4) | 35 (46.6) |
| Follow-up, y, mean (SD) | 3.9 (1.5) | 4 (1.4) | 3.7 (1.5) |
| [$^{18}$F]Florbetapir SUVR, mean (SD) | .77 (.11) | .70 (.04) | .89 (.10) |
| CSF p-tau<sub>181</sub>, pg/mL <sup>a</sup>, mean (SD) | 22.58 (9.37) | 20.34 (6.99) | 26.45 (11.52) |
| PACC, mean (SD) | -.14 (2.6) | .03 (2.6) | -.48 (2.6) |
| ADNI-EF, mean (SD) | .85 (.82) | .96 (.84) | .63 (.74) |
| ADNI-Mem, mean (SD) | 1.06 (.56) | 1.11 (.56) | .98 (.53) |
| VRFs <sup>b</sup>, mean (SD) | 1.55 (1.15) | 1.47 (1.12) | 1.70 (1.20) |
| GDS, mean (SD) | .88 (1.11) | .95 (1.18) | .74 (.97) |

Abbreviations: SD, standard deviation; y, years; APOE ε4, apolipoprotein E ε4; SUVR, standardized uptake value ratio; CSF, cerebrospinal fluid; p-tau<sub>181</sub>, hyperphosphorylated tau; Aβ , amyloid-β ; PACC, Preclinical Alzheimer's Cognitive Composite; ADNI-EF, Alzheimer's
Disease Neuroimaging Initiative composite score for executive function; ADNI-Mem, Alzheimer's Disease Neuroimaging Initiative composite score for memory function; VRFs, vascular risk factors; GDS, Geriatric Depression Scale.
<sup>a</sup>Total of 193 subjects (122 Aβ-negative and 71 Aβ-positive).
<sup>b</sup>Represents the sum of VRFs.

### ADHD-PRS is associated with a persistent executive function deficit from baseline

We observed a significant main effect of ADHD-PRS on ADNI-EF (β = -.09, 95% CI = -.18 to -.008, p-value = .03), suggesting that higher ADHD-PRS was related to a persistent executive function deficit in all time points including baseline (Figure 1A). No significant effect was observed for ADHD-PRS in the model with PACC global cognitive composite as the outcome variable (β = -.12, 95% CI = -.44 to 19, p-value = .45; Figure 1B) or in the model containing ADNI-Mem as the outcome variable (β = .01, 95% CI = -.05 to .08, p-value = .63; Figure 1C).

**Figure 1.**
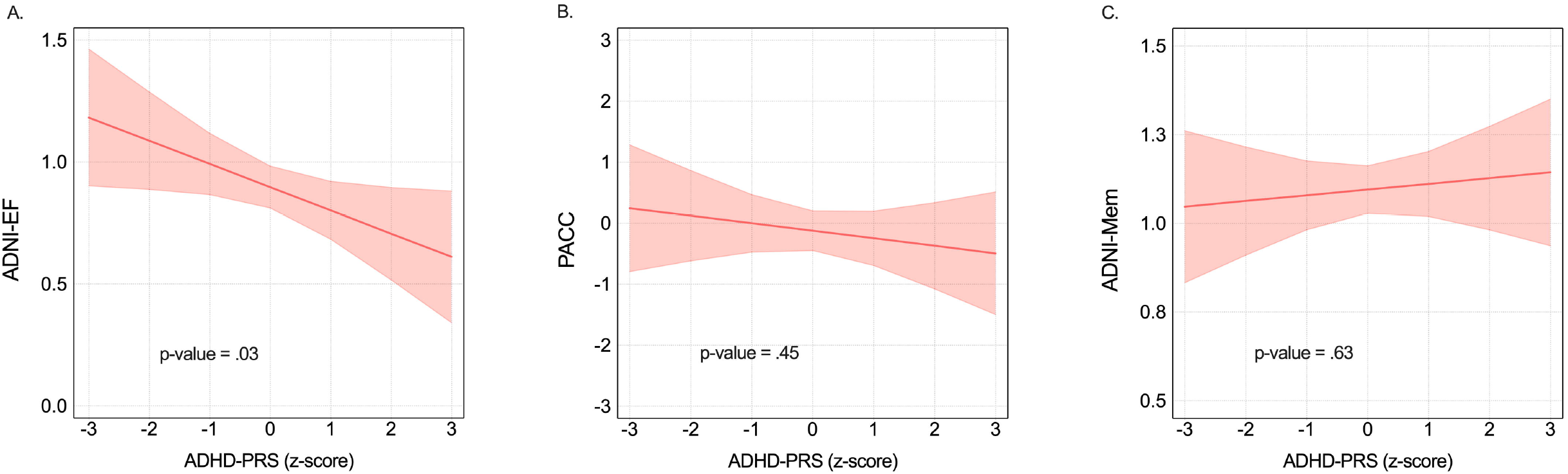
ADHD-PRS is associated with a persistent deficit in executive function from the baseline visit. Figure 1 shows the association between the main effect of ADHD-PRS and cognitive functions accounting for sex, age at baseline, *APOE* ε4 carriership status (carriers vs. non-carriers), years of education, and ancestry (using the first seven PCs to account for ancestry differences in genetics structure that could bias the results). P-values represent the main effect of ADHD-PRS on cognitive function including all visits. Lines and shaded areas reflect the estimated marginal means from mixed effect models analyses ± 95% CI. The model is described in the Supplementary Material (model 1). Abbreviations: ADHD-PRS, Attention-Deficit/Hyperactivity Disorder polygenic risk score; ADNI-EF, Alzheimer’s Disease Neuroimaging Initiative composite score for executive function; PACC, Preclinical Alzheimer’s Cognitive Composite; ADNI-Mem, Alzheimer’s Disease Neuroimaging Initiative composite score for memory function; *APOE* ε4, apolipoprotein E ε4; PCs, principal components; CI, confidence interval.

### ADHD-PRS associates with longitudinal cognitive decline

We observed a significant interaction between ADHD-PRS and time on PACC, demonstrating that higher ADHD-PRS was associated with a higher decline in general cognitive performance over 6 years (ADHD-PRS x time; β = -.10, 95% CI = -.16 to -.03 p-value = .003; Figure 2A) with a Cohen’s *f*^2^ of .21, indicating a medium effect size. As a comparison, the Cohen’s *f*^2^ for baseline [^18^F]florbetapir Aβ-PET, a well-established predictor of cognitive decline in AD, was .48, indicating a large effect size. Similar findings were observed for ADNI-Mem (ADHD-PRS x time; β = -.01, 95% CI = -.02 to -.002, p-value = .01; Figure 2B), indicating that higher ADHD-PRS was related to a progressive decline in memory function. No longitudinal effects were observed for ADNI-EF (ADHD-PRS x time; β = -.003, 95% CI = -.01 to .01, p-value = .62; Figure 2C).

**Figure 2.**
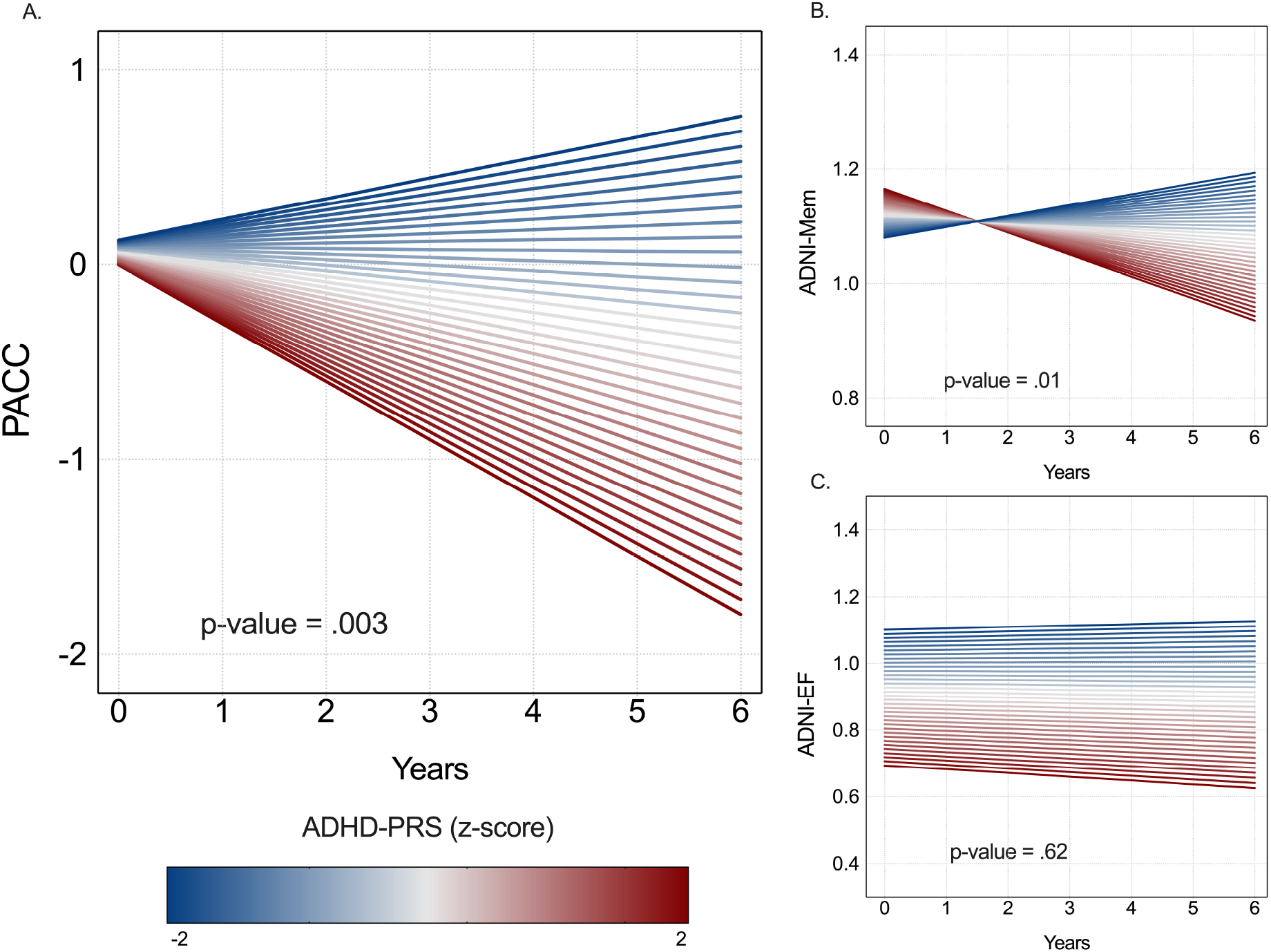
Higher ADHD-PRS is associated with longitudinal cognitive decline over 6 years in CU older adults. Figure 2 shows that higher ADHD-PRS was associated with decreased performance on general cognitive performance (PACC) and memory (ADNI-Mem) over time. The p-values represent the effect of ADHD-PRS on cognition over time. Lines reflect the estimated marginal means from mixed effect models analyses. The model is described in the Supplementary Material (model 2). Abbreviations: ADHD-PRS, Attention-Deficit/Hyperactivity Disorder polygenic risk score; CU, cognitively unimpaired; PACC, Preclinical Alzheimer’s Cognitive Composite; ADNI-Mem, Alzheimer’s Disease Neuroimaging Initiative composite score for memory function; ADNI-EF, Alzheimer’s Disease Neuroimaging Initiative composite score for executive function; PCs, principal components.

### ADHD-PRS and Aβ show an interaction in longitudinal cognitive decline

We observed a significant interaction between ADHD-PRS and baseline Aβ-PET on worsening cognitive performance, indicating that the association between higher ADHD-PRS and decreased PACC scores over time was present in Aβ-positive but not in Aβ-negative individuals (ADHD-PRS x time x baseline Aβ-PET; β = -.17, 95% CI = -.31 to -.02, p-value = .01; Figure 3A). Similar findings were obtained for ADNI-Mem (ADHD-PRS x time x baseline Aβ-PET; β = -.02, 95% CI = -.05 to -.0001, p-value = .04; Figure 3B), but not ADNI-EF (ADHD-PRS x time x baseline Aβ-PET; β = .001, 95% CI = -.03 to .03, p-value = .92; Figure 3C).

**Figure 3.**
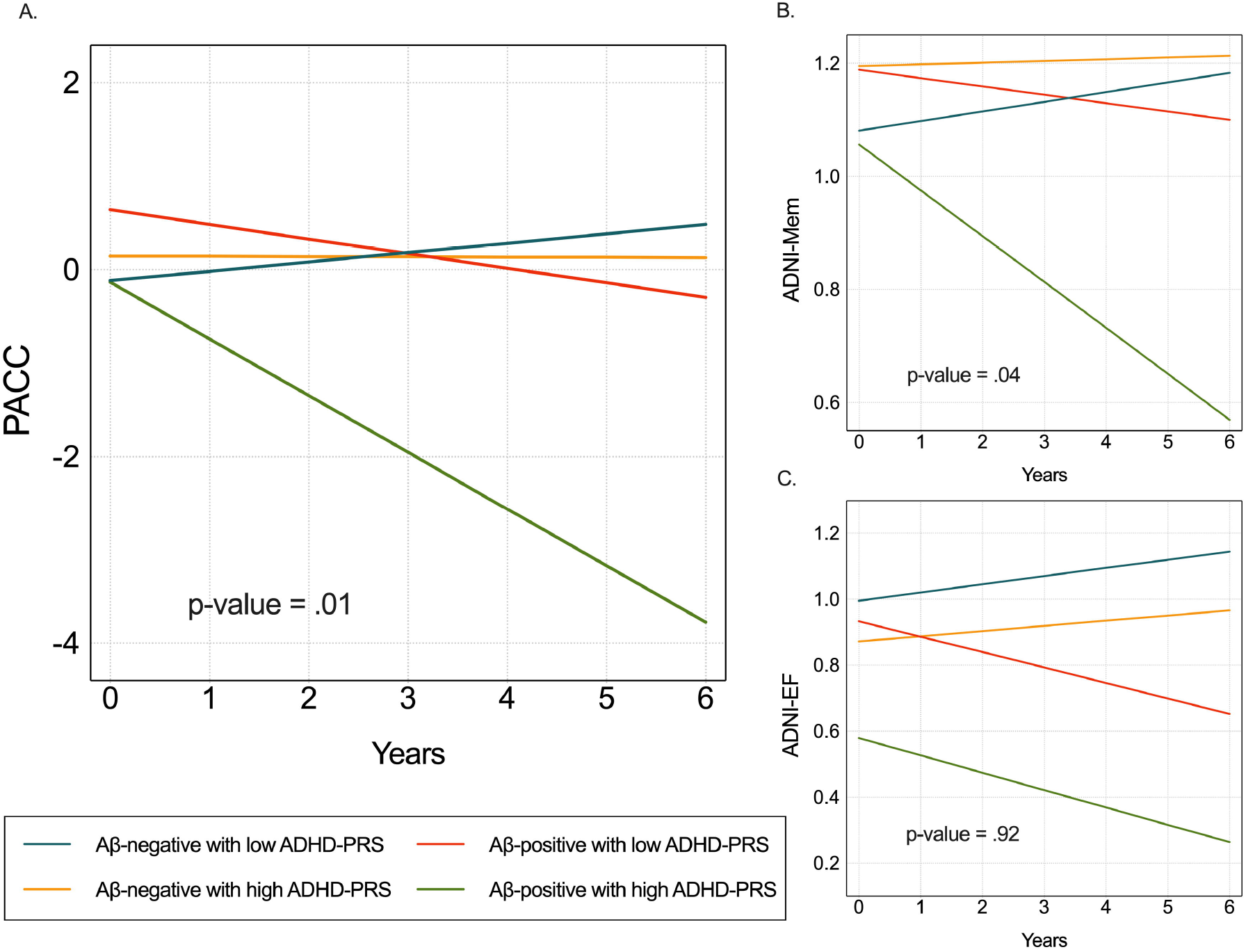
Aβ-positivity and high ADHD-PRS potentiated longitudinal cognitive impairment in CU older adults. Figure 3 shows that higher ADHD-PRS was associated with decreased performance on general cognitive performance (PACC) and memory (ADNI-Mem) over time only in Aβ-positive individuals. The p-values represent the significance of the interaction term between ADHD-PRS and baseline Aβ burden on cognition over time. Lines reflect the estimated marginal means from mixed effect models analyses. Low ADHD-PRS includes z-score of −1, and high ADHD-PRS includes z-score of 1. The model is described in the Supplementary Material (model 3). Abbreviations: ADHD-PRS, Attention-Deficit/Hyperactivity Disorder polygenic risk score; CU, cognitively unimpaired; PACC, Preclinical Alzheimer’s Cognitive Composite; ADNI-Mem, Alzheimer’s Disease Neuroimaging Initiative composite score for memory function; ADNI-EF, Alzheimer’s Disease Neuroimaging Initiative composite score for executive function; Aβ, amyloid-β; PCs, principal components.

### ADHD-PRS associates with forthcoming tau pathology and brain atrophy in Aβ-positive individuals

Higher ADHD-PRS was highly associated with increased CSF p-tau_181_ over time in Aβ-positive individuals (ADHD-PRS x time; β = .05, 95% CI = .01 to .08, p-value = .003, Figure 4B). On the other hand, no significant association was observed for Aβ-negative individuals (ADHD-PRS x time; β = -.003, 95% CI = -.02 to .01, p-value = .70, Figure 4A). Similarly, ADHD-PRS was associated with longitudinal neurodegeneration as measured by reduction of GM density in the superior frontal gyrus and supramarginal gyrus (Figure 5) in Aβ-positive individuals.

**Figure 4.**
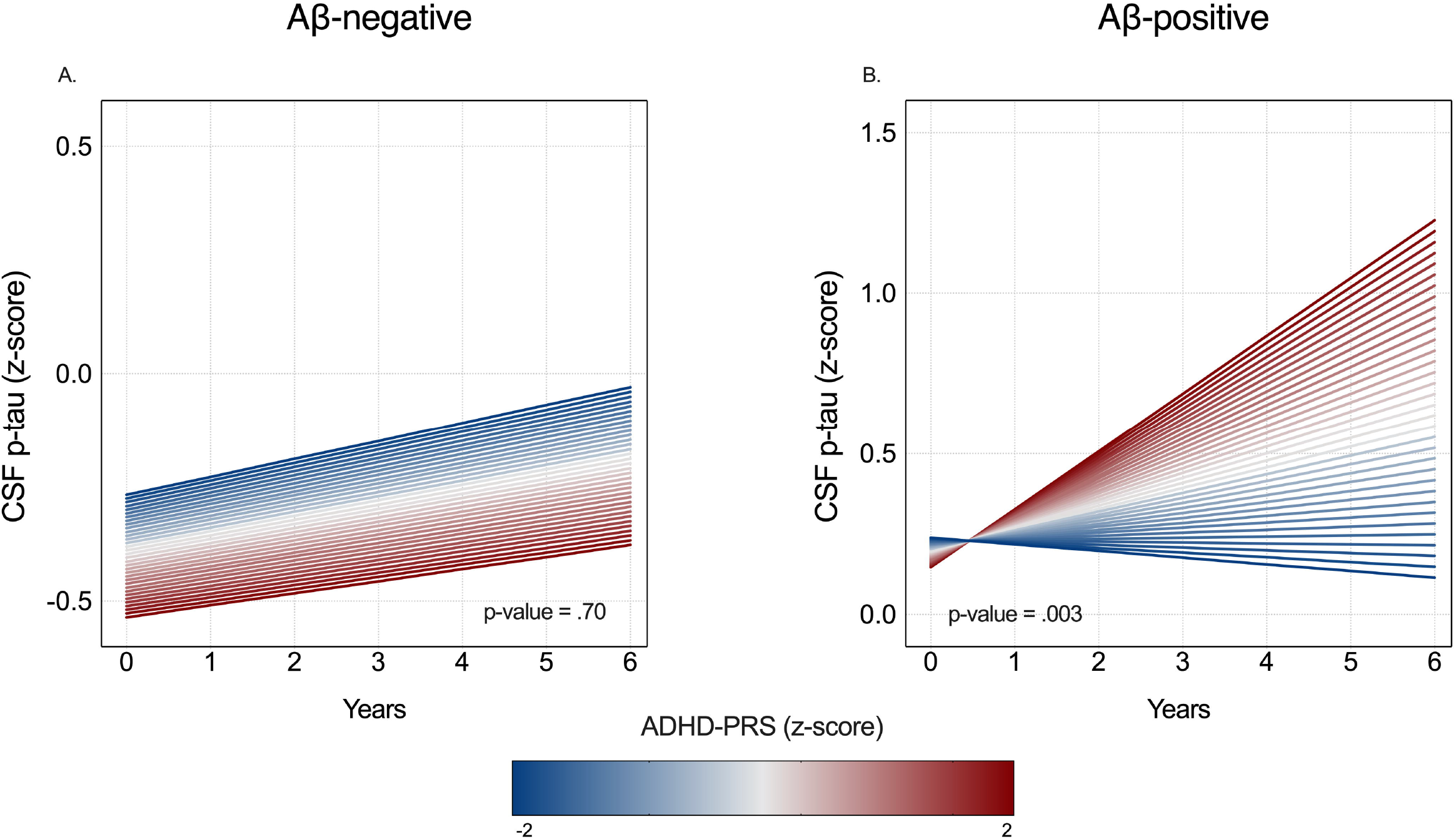
ADHD-PRS is associated with the development of tau pathology over 6 years only in Aβ-positive individuals. Figure 4 shows that ADHD-PRS was associated with increased CSF p-tau_181_ in Aβ-positive but not in Aβ-negative individuals over a 6-year time frame. The p-values represent the significance of the interaction term between ADHD-PRS and baseline Aβ burden on CSF p-tau over time. Lines reflect the estimated marginal means from the mixed effect models analyses. The model is described in the Supplementary Material (model 4). Abbreviations: CSF, cerebrospinal fluid; p-tau, tau phosphorylated at threonine 181; Aβ, amyloid-β; ADHD-PRS, Attention-Deficit/Hyperactivity Disorder polygenic risk score.

**Figure 5.**
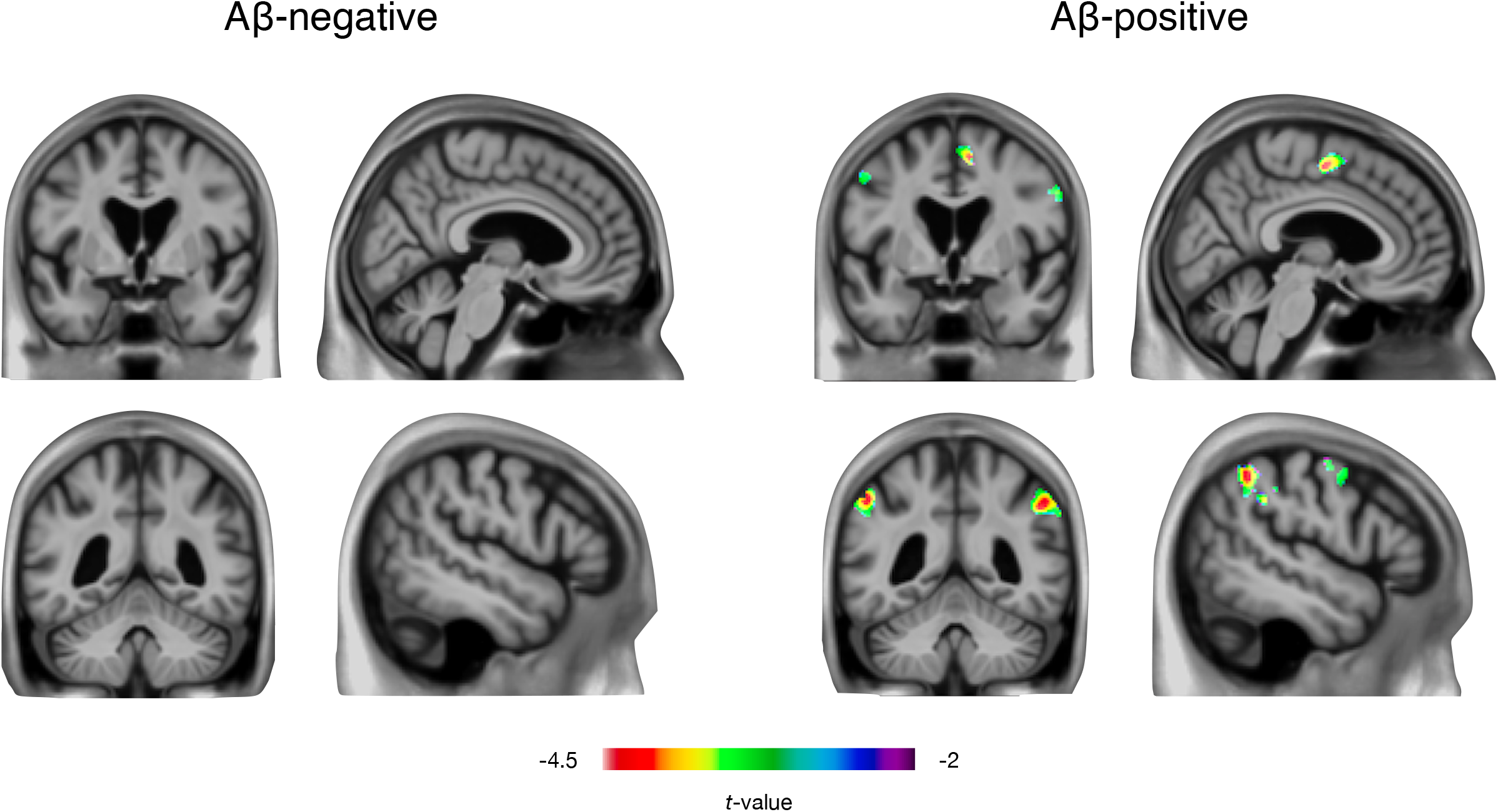
ADHD-PRS is associated with longitudinal brain atrophy over 6 years in the frontal and parietal cortices of CU Aβ-positive older individuals. Figure 5 shows that ADHD-PRS was associated with a longitudinal decrease in GM density in the superior frontal gyrus and supramarginal gyrus of CU Aβ-positive older individuals. The t-statistical parametric images show the result of the voxel-wise linear mixed-effects model testing the interaction between ADHD-PRS and time on GM. We used RFT to correct the results for multiple comparisons at a threshold of *P* < .001. The model is described in the Supplementary Material (model 5). Abbreviations: ADHD-PRS, Attention-Deficit/Hyperactivity Disorder polygenic risk score; CU, cognitively unimpaired; GM, gray matter; Aβ, amyloid-β; RFT, random field theory.

### Sensitivity analyses

The aforementioned findings were replicated using most ADHD-PRS thresholds, supporting the robustness of our results (Table S2). In addition, similar findings were obtained by adjusting for possible confounders such as VRFs and depression symptoms (Table S3 and Table S4).

## Discussion

This study aimed to determine whether ADHD-PRS was associated with longitudinal cognitive impairment in CU older adults and whether this association was related to the core markers of AD pathology. For the first time, we described that higher ADHD-PRS was associated with progressive longitudinal cognitive decline, particularly in memory function. Furthermore, cognitive decline was mostly observed in Aβ-positive individuals, suggesting that individuals carrying a genetic liability for ADHD are characterized by cognitive susceptibility to the presence of Aβ pathology. Finally, in Aβ-positive individuals, higher ADHD-PRS was associated with longitudinal increases in CSF p-tau_181_ and brain atrophy in frontal and parietal brain regions. These findings suggest that the genetic liability to ADHD increases the susceptibility to cognitive decline, tau pathology, and neurodegeneration in the presence of Aβ pathology in the brain of older individuals.

Our results corroborate previous studies showing deficits in executive function among older adults with ADHD (3). We observed that higher genetic liability for ADHD was associated with executive function deficits, which remained relatively constant over time. These results are unsurprising and consistent with previous literature showing an association between higher ADHD-PRS and decreased executive function during childhood (9). Importantly, our study provides unique evidence that higher ADHD-PRS was associated with progressive cognitive decline, predominantly in the memory domain. Since the prototypical clinical phenotype of AD is progressive amnestic symptoms (23), our results support previous epidemiological findings demonstrating that ADHD might be a risk factor for cognitive decline, potentially leading to MCI and dementia syndromes due to AD (6–8).

We found that future cognitive decline in older adults is associated with the presence of both high ADHD-PRS and brain Aβ pathology. Specifically, ADHD-PRS potentiated the effects of Aβ on longitudinal clinical and pathophysiological progressions in our older population. It is postulated that abnormal Aβ deposition triggers a cascade of events leading to AD progression (23). Although brain Aβ load is associated with AD-related cognitive decline (25), Aβ pathology alone seems not to be not sufficient to cause it (25). For example, it is well established that around 30% of CU individuals older than 55 years of age present brain Aβ pathology and that a large portion of these individuals remains cognitively intact during their lives (26). This supports that the association between Aβ accumulation and cognitive deterioration depends on patients’ intrinsic resilience and susceptibility mechanisms. Together, the results above indicate that the genetic liability to ADHD plays a role in increasing the susceptibility to the harmful effects of Aβ in the human brain.

A widely held view in the AD field posits that Aβ triggers the spread of tau pathology, leading to neurodegeneration and cognitive impairment (25). In line with this hypothesis, our study showed that the association between ADHD-PRS and cognitive decline was accompanied by a longitudinal increase in CSF levels of p-tau_181_, a well-validated marker of brain tau pathology (27). Our findings suggest that ADHD-PRS is related to both tau deposition and cognitive decline in Aβ-positive individuals, highlighting the genetic liability for ADHD as a relevant factor influencing AD progression in the presence of Aβ pathology.

We showed that higher ADHD-PRS was associated with brain atrophy in frontal and parietal brain regions in Aβ-positive individuals. Specifically, decreased GM density over time was observed in the superior frontal gyrus and supramarginal gyrus. Previous AD studies have shown that brain atrophy closely correlates with tau deposition and cognitive deficits, being a well-established marker of disease progression (28). In CU populations, AD-related brain atrophy has been reported predominantly in the medial temporal cortex. In contrast, reduced cortical thickness in regions such as the superior frontal gyrus and supramarginal gyrus is present in later stages of AD (28). Interestingly, atrophy in parietal and frontal cortices has been demonstrated in middle-aged (29) and older adults (30) with ADHD. These results suggest that while in our study ADHD-PRS-related atrophy in the superior and supramarginal gyrus was Aβ pathology-dependent, it recapitulated regions showing atrophy in ADHD rather than early AD. This supports the notion that the combination of underlying ADHD-related vulnerability with Aβ pathology is a factor associated with cognitive dysfunction in older adults.

This study should be viewed in light of some limitations. First, our sample did not have a detailed clinical assessment for ADHD diagnosis. Since ADHD is a relatively recent diagnostic category, older adults are unlikely to have received the diagnosis as children (1). Thus, ADHD-PRS in our results may identify either asymptomatic older adults with genetic susceptibility to ADHD and/or patients with undiagnosed ADHD. In addition, the fact that neuropsychiatric conditions such as depression, bipolar disorder, or schizophrenia were not included in the ADNI cohort further limits the external validity of our results. The sample included in this study likely represents a less affected population when compared to the general ADHD population treated in clinical centers, where psychiatric comorbidities are the rule rather than the exception (1). The population used to generate ADHD-PRS (12) and our study population were composed almost exclusively of white participants. Therefore, future studies in ADHD and AD should focus on enrolling more diverse populations. Finally, no adjustments were made to control type I error, and consequently, the analyses should be considered exploratory.

## Conclusion

To conclude, our results suggest that ADHD-PRS can be used to inform the risk of cognitive decline in Aβ-positive CU older adults. Since ADHD-PRS was associated with cognitive decline in our entire population, ADHD-PRS may also be used to predict cognitive deterioration in the absence of AD biomarkers. Importantly, ADHD-PRS was associated with longitudinal CSF p-tau hyperphosphorylation and brain atrophy in frontoparietal but not temporal regions in Aβ-positive individuals, suggesting that an ADHD-related brain susceptibility to the harmful effects of Aβ plays a role in the early development of AD in genetically vulnerable patients.

## Data Availability

All data produced in the present study are available upon reasonable request to the authors.

## Acknowledgments

DTL is supported by a CNPq postdoctoral fellowship (#154116/2018-1), and supported by a NARSAD Young Investigator Grant from the Brain & Behavior Research Foundation (#29486). JPF-S receives financial support from CAPES (#88887.627297/2021-00). CT receives funding from Faculty of Medicine McGill and IPN McGill. PCLF is supported by the Alzheimer’s Association (#AARFD-22-923814). WSB is supported by CAPES (#88887.372371/2019-00 and #88887.596742/2020-00). TKK is funded by the Swedish Research Council’s career establishment fellowship (#2021-03244), the Alzheimer’s Association Research Fellowship (#850325), the BrightFocus Foundation (#A2020812F), the International Society for Neurochemistry’s Career Development Grant, the Swedish Alzheimer Foundation (Alzheimerfonden; #AF-930627), the Swedish Brain Foundation (Hjärnfonden; #FO2020-0240), the Swedish Dementia Foundation (Demensförbundet), the Swedish Parkinson Foundation (Parkinsonfonden), Gamla Tjänarinnor Foundation, the Aina (Ann) Wallströms and Mary-Ann Sjöbloms Foundation, the Agneta Prytz-Folkes & Gösta Folkes Foundation (#2020-00124), the Gun and Bertil Stohnes Foundation, and the Anna Lisa and Brother Björnsson’s Foundation. ERZ receives financial support from CNPq (#435642/2018-9 and #312410/2018-2), Instituto Serrapilheira (#Serra-1912-31365), Brazilian National Institute of Science and Technology in Excitotoxicity and Neuroprotection (#465671/2014-4), FAPERGS/MS/CNPq/SESRS–PPSUS (#30786.434.24734.231120170), ARD/FAPERGS (#54392.632.30451.05032021), and Alzheimer’s Association (#AARGD-21-850670). TAP is supported by the NIH (#R01AG075336 and #R01AG073267) and the Alzheimer’s Association (#AACSF-20-648075).

## References

1. Faraone S V, Banaschewski T, Coghill D, Zheng Y, Biederman J, Bellgrove MA, et al. The World Federation of ADHD International Consensus Statement: 208 Evidence-based Conclusions about the Disorder. Neurosci Biobehav Rev [Internet]. 2021; Available from: https://www.sciencedirect.com/science/article/pii/S014976342100049X

2. Sibley MH, Arnold LE, Swanson JM, Hechtman LT, Kennedy TM, Owens E, et al. Variable Patterns of Remission From ADHD in the Multimodal Treatment Study of ADHD. Am J Psychiatry. 2021;appi.ajp.2021.2.

3. Kooij JJS, Michielsen M, Kruithof H, Bijlenga D. ADHD in old age: a review of the literature and proposal for assessment and treatment. Expert Rev Neurother. 2016;16(12):1371–81.

4. Dobrosavljevic M, Solares C, Cortese S, Andershed H, Larsson H. Prevalence of attention-deficit/hyperactivity disorder in older adults: A systematic review and meta-analysis. Neurosci Biobehav Rev [Internet]. 2020;118(February):282–9. Available from: https://doi.org/10.1016/j.neubiorev.2020.07.042

5. Callahan BL, Bierstone D, Stuss DT, Black SE. Adult ADHD: Risk factor for dementia or phenotypic mimic? Front Aging Neurosci. 2017;9(AUG):1–15.

6. Du Rietz E, Brikell I, Butwicka A, Leone M, Chang Z, Cortese S, et al. Mapping phenotypic and aetiological associations between ADHD and physical conditions in adulthood in Sweden: a genetically informed register study. The lancet Psychiatry. 2021 Jul;

7. Zhang L, Du Rietz E, Kuja-Halkola R, Dobrosavljevic M, Johnell K, Pedersen NL, et al. Attention-deficit/hyperactivity disorder and Alzheimer’s disease and any dementia: A multi-generation cohort study in Sweden. Alzheimer’s Dement. 2021;(November 2020):1–9.

8. Dobrosavljevic M, Zhang L, Garcia-Argibay M, Du Rietz E, Andershed H, Chang Z, et al. Attention-deficit/hyperactivity disorder as a risk factor for dementia and mild cognitive impairment: A population-based register study. Eur Psychiatry [Internet]. 2021/12/20. 2022;65(1):e3. Available from: https://www.cambridge.org/core/article/attentiondeficithyperactivity-disorder-as-a-risk-factor-for-dementia-and-mild-cognitive-impairment-a-populationbased-register-study/E289DAD7BE1BFB781389CF369EF7ECB3

9. Ronald A, de Bode N, Polderman TJC. Systematic Review: How the Attention-Deficit/Hyperactivity Disorder Polygenic Risk Score Adds to Our Understanding of ADHD and Associated Traits. J Am Acad Child Adolesc Psychiatry. 2021 Feb;

10. Petersen RC, Aisen PS, Beckett LA, Donohue MC, Gamst AC, Harvey DJ, et al. Alzheimer’s Disease Neuroimaging Initiative (ADNI): clinical characterization. Neurology. 2010 Jan;74(3):201–9.

11. Zettergren A, Lord J, Ashton NJ, Benedet AL, Karikari TK, Lantero Rodriguez J, et al. Association between polygenic risk score of Alzheimer’s disease and plasma phosphorylated tau in individuals from the Alzheimer’s Disease Neuroimaging Initiative. Alzheimers Res Ther [Internet]. 2021;13(1):17. Available from: https://doi.org/10.1186/s13195-020-00754-8

12. Demontis D, Walters RK, Martin J, Mattheisen M, Als TD, Agerbo E, et al. Discovery of the first genome-wide significant risk loci for attention deficit/hyperactivity disorder. Nat Genet. 2019 Jan;51(1):63–75.

13. Euesden J, Lewis CM, O’Reilly PF. PRSice: Polygenic Risk Score software. Bioinformatics. 2015 May;31(9):1466–8.

14. Chang CC, Chow CC, Tellier LC, Vattikuti S, Purcell SM, Lee JJ. Second-generation PLINK: rising to the challenge of larger and richer datasets. Gigascience. 2015;4:7.

15. Donohue MC, Sperling RA, Salmon DP, Rentz DM, Raman R, Thomas RG, et al. The preclinical Alzheimer cognitive composite: Measuring amyloid-related decline. JAMA Neurol. 2014;71(8):961–70.

16. Gibbons LE, Carle AC, Mackin RS, Harvey D, Mukherjee S, Insel P, et al. A composite score for executive functioning, validated in Alzheimer’s Disease Neuroimaging Initiative (ADNI) participants with baseline mild cognitive impairment. Brain Imaging Behav. 2012 Dec;6(4):517–27.

17. Crane PK, Carle A, Gibbons LE, Insel P, Mackin RS, Gross A, et al. Development and assessment of a composite score for memory in the Alzheimer’s Disease Neuroimaging Initiative (ADNI). Brain Imaging Behav. 2012 Dec;6(4):502–16.

18. Jack CRJ, Bernstein MA, Fox NC, Thompson P, Alexander G, Harvey D, et al. The Alzheimer’s Disease Neuroimaging Initiative (ADNI): MRI methods. J Magn Reson Imaging. 2008 Apr;27(4):685–91.

19. Landau SM, Mintun MA, Joshi AD, Koeppe RA, Petersen RC, Aisen PS, et al. Amyloid deposition, hypometabolism, and longitudinal cognitive decline. Ann Neurol. 2012;72(4):578–86.

20. Mathotaarachchi S, Wang S, Shin M, Pascoal TA, Benedet AL, Kang MS, et al. VoxelStats: A MATLAB Package for Multi-Modal Voxel-Wise Brain Image Analysis [Internet]. Vol. 10, Frontiers in Neuroinformatics 2016. Available from: https://www.frontiersin.org/article/10.3389/fninf.2016.00020

21. Selya A, Rose J, Dierker L, Hedeker D, Mermelstein R. A Practical Guide to Calculating Cohen’s f2, a Measure of Local Effect Size, from PROC MIXED [Internet]. Vol. 3, Frontiers in Psychology 2012. Available from: https://www.frontiersin.org/article/10.3389/fpsyg.2012.00111

22. Keller MC. Gene × environment interaction studies have not properly controlled for potential confounders: the problem and the (simple) solution. Biol Psychiatry. 2014 Jan;75(1):18–24.

23. Knopman DS, Amieva H, Petersen RC, Chételat G, Holtzman DM, Hyman BT, et al. Alzheimer disease. Nat Rev Dis Prim [Internet]. 2021;7(1):33. Available from: https://doi.org/10.1038/s41572-021-00269-y

24. Ferrari-Souza JP, Brum WS, Hauschild LA, Da Ros LU, Lukasewicz Ferreira PC, Bellaver B, et al. Vascular risk burden is a key player in the early progression of Alzheimer’s disease. medRxiv [Internet]. 2021 Jan 1;2021.12.18.21267994. Available from: http://medrxiv.org/content/early/2021/12/19/2021.12.18.21267994.abstract

25. Long JM, Holtzman DM. Alzheimer Disease: An Update on Pathobiology and Treatment Strategies. Cell [Internet]. 2019;179(2):312–39. Available from: https://doi.org/10.1016/j.cell.2019.09.001

26. Jansen WJ, Janssen O, Tijms BM, Vos SJB, Ossenkoppele R, Visser PJ, et al. Prevalence Estimates of Amyloid Abnormality Across the Alzheimer Disease Clinical Spectrum. JAMA Neurol [Internet]. 2022 Jan 31; Available from: https://doi.org/10.1001/jamaneurol.2021.5216

27. La Joie R, Bejanin A, Fagan AM, Ayakta N, Baker SL, Bourakova V, et al. Associations between [(18)F]AV1451 tau PET and CSF measures of tau pathology in a clinical sample. Neurology. 2018 Jan;90(4):e282–90.

28. Frisoni GB, Fox NC, Jack Jr CR, Scheltens P, Thompson PM. The clinical use of structural MRI in Alzheimer disease. Nat Rev Neurol [Internet]. 2010 Feb;6(2):67–77. Available from: https://pubmed.ncbi.nlm.nih.gov/20139996

29. Moreno-Alcázar A, Ramos-Quiroga JA, Radua J, Salavert J, Palomar G, Bosch R, et al. Brain abnormalities in adults with Attention Deficit Hyperactivity Disorder revealed by voxel-based morphometry. Psychiatry Res Neuroimaging [Internet]. 2016;254:41– 7. Available from: https://www.sciencedirect.com/science/article/pii/S0925492715301815

30. Klein M, Souza-Duran FL, Menezes AKPM, Alves TM, Busatto G, Louzã MR. Gray Matter Volume in Elderly adults With ADHD: Associations of Symptoms and Comorbidities With Brain Structures. J Atten Disord. 2021 Apr;25(6):829–38.

