## Supplementary material for "Genetic Risk for Attention-Deficit/Hyperactivity Disorder Predicts Cognitive Decline and Development of Alzheimer’s Disease Pathophysiology in Cognitively Unimpaired Older Adults"

SUPPLEMENTARY MATERIAL: TABLE OF CONTENTS

### **Supplementary Methods**

#### **Vascular burden**

Vascular risk factors (VRFs) were assessed using a composite score, as previously described (1). Briefly, we calculated the score based on the presence or absence of the following conditions: (1) cardiovascular disease [coronary artery disease (myocardial infarction, angina, stent placement, angioplasty, coronary artery bypass graft, coronary insufficiency), heart failure, or intermittent claudication]; (2) hypertension; (3) diabetes mellitus; (4) hyperlipidemia; (5) stroke or transient ischemic attack (TIA); (6) smoking (ever or never); (7) atrial fibrillation; and (8) left ventricular hypertrophy. The score was obtained as the sum of VRFs (ranging from zero to eight). An elevated VRF burden was defined as a vascular composite score equal to or higher than two (1).

#### **Statistical models**

In the statistical models described below:

i. *β*_0_ represents the intercept;

ii. ${ADHD-PRS}_{i}$ represents the ADHD-PRS for subject *i*;

iii. ${Time}_{ij}$ represents the effect of time, fitted as a continuous measure in years;

iv. ${Sex}_{i}$ is the dummy variable for sex (1 = male, 0 = female);

iv. ${Age}_{i}$represents years of age for subject *i* at baseline;

v. ${APOE \varepsilon4}_{i}$ represents the dummy variable for apolipoprotein E ε4 (*APOE* ε4) carriership status (1 = positive, 0 = negative);

vi. ${Years of education}_{i}$ represents years of formal education for subject *i* at baseline, fitted as a continuous measure in years;

vii. ${Ancestry}_{i}$ represents the first seven principal components, which were included as recommended for ADNI datasets (2) to account for any ancestry differences in genetic structure that could bias the results;

viii. ${A\beta-PET}_{i}$ represents the dummy variable for Aβ positivity (1 = Aβ-positive, 0 = Aβ-negative);

ix. ${VRF}_{i}$ represents the dummy variable for VRF positivity (1 = positive, vascular composite score ≥ 2, 0 = negative, vascular composite score ≤ 1);

x. ${GDS}_{i}$ represents the GDS scores at baseline, fitted as a continuous measure;

xi. $\upsilon_{0i}$ is the subject-specific variation from the average intercept effect, and $\varepsilon_{ij}$ is the random error term at the *j^th^* time point for subject *I.*

* Models 4 and 5 were conducted separately on individuals Aβ positive and negative.

Model 1:

$$\Delta Cognitive function=\beta_{0}+\beta_{1}{ADHD-PRS}_{i}+\beta_{2}{Time}_{ij}+\beta_{3}{Sex}_{i}+\beta_{4}{Age}_{i}+\beta_{5}{APOE \varepsilon4}_{i}+\beta_{6}{Years of education}_{i}+\beta_{7}{Ancestry}_{i}+\upsilon_{0i}+\varepsilon_{ij}$$

Model 2:

$$\Delta Cognitive function=\beta_{0}+\beta_{1}{ADHD-PRS}_{i} X {Time}_{ij}+\beta_{1}{ADHD-PRS}_{i}+\beta_{2}{Time}_{ij}+\beta_{2}{ADHD-PRS}_{i} X {Sex}_{i}+\beta_{3}{Time}_{ij} X {Sex}_{i}+\beta_{4}{Sex}_{i}+\beta_{5}{ADHD-PRS}_{i} X {Age}_{i}+\beta_{6}{Time}_{ij} X {Age}_{i}+\beta_{7}{Age}_{i}+\beta_{8}{ADHD-PRS}_{i} X {APOE \varepsilon4}_{i}+{\beta_{9}{Time}_{ij} X {APOE \varepsilon4}_{i}+\beta}_{10}{APOE \varepsilon4}_{i}+\beta_{11}{ADHD-PRS}_{i} X {Years of education}_{i}+\beta_{12}{Time}_{ij} X {Years of education}_{i}+\beta_{13}{Years of education}_{i}+\beta_{14}{Ancestry}_{i}+\upsilon_{0i}+\varepsilon_{ij}$$

Model 3:

$$\Delta Cognitive function=\beta_{0}+\beta_{1}{ADHD-PRS}_{i} X {Time}_{ij}X {A\beta-PET}_{i}+\beta_{2}{ADHD-PRS}_{i}+\beta_{3}{Time}_{ij}+{\beta_{4}A\beta-PET}_{i}+\beta_{5}{ADHD-PRS}_{i} X {Sex}_{i}+\beta_{6}{Time}_{ij} X {Sex}_{i}+\beta_{7}{A\beta-PET}_{i} X {Sex}_{i}+\beta_{8}{Sex}_{i}+\beta_{9}{ADHD-PRS}_{i} X {Age}_{i}+\beta_{10}{Time}_{ij} X {Age}_{i}+{\beta_{11}{A\beta-PET}_{i} X {Age}_{i}+\beta}_{7}{Age}_{i}+\beta_{12}{ADHD-PRS}_{i} X {APOE \varepsilon4}_{i}+{\beta_{13}{Time}_{ij} X {APOE \varepsilon4}_{i}+\beta_{14}{A\beta-PET}_{i} X {APOE \varepsilon4}_{i}+\beta}_{15}{APOE \varepsilon4}_{i}+\beta_{16}{ADHD-PRS}_{i} X {Years of education}_{i}+\beta_{17}{Time}_{ij} X {Years of education}_{i}+\beta_{18}{A\beta-PET}_{i} X {Years of education}_{i}+\beta_{19}{Years of education}_{i}+\beta_{20}{Ancestry}_{i}+\upsilon_{0i}+\varepsilon_{ij}$$

Model 4:

$$\Delta{CSF p-tau}_{181}=\beta_{0}+\beta_{1}{ADHD-PRS}_{i} X {Time}_{ij}+\beta_{1}{ADHD-PRS}_{i}+\beta_{2}{Time}_{ij}+\beta_{2}{ADHD-PRS}_{i} X {Sex}_{i}+\beta_{3}{Time}_{ij} X {Sex}_{i}+\beta_{4}{Sex}_{i}+\beta_{5}{ADHD-PRS}_{i} X {Age}_{i}+\beta_{6}{Time}_{ij} X {Age}_{i}+\beta_{7}{Age}_{i}+\beta_{8}{ADHD-PRS}_{i} X {APOE \varepsilon4}_{i}+{\beta_{9}{Time}_{ij} X {APOE \varepsilon4}_{i}+\beta}_{10}{APOE \varepsilon4}_{i}+\beta_{11}{ADHD-PRS}_{i} X {Years of education}_{i}+\beta_{12}{Time}_{ij} X {Years of education}_{i}+\beta_{13}{Years of education}_{i}+\beta_{14}{Ancestry}_{i}+\upsilon_{0i}+\varepsilon_{ij}$$

Model 5:

$$\Delta VBM=\beta_{0}+\beta_{1}{ADHD-PRS}_{i} X {Time}_{ij}+\beta_{1}{ADHD-PRS}_{i}+\beta_{2}{Time}_{ij}+\beta_{2}{ADHD-PRS}_{i} X {Sex}_{i}+\beta_{3}{Time}_{ij} X {Sex}_{i}+\beta_{4}{Sex}_{i}+\beta_{5}{ADHD-PRS}_{i} X {Age}_{i}+\beta_{6}{Time}_{ij} X {Age}_{i}+\beta_{7}{Age}_{i}+\beta_{8}{ADHD-PRS}_{i} X {APOE \varepsilon4}_{i}+{\beta_{9}{Time}_{ij} X {APOE \varepsilon4}_{i}+\beta}_{10}{APOE \varepsilon4}_{i}+\beta_{11}{ADHD-PRS}_{i} X {Years of education}_{i}+\beta_{12}{Time}_{ij} X {Years of education}_{i}+\beta_{13}{Years of education}_{i}+\beta_{14}{Ancestry}_{i}+\upsilon_{0i}+\varepsilon_{ij}$$

Model 6:

$$\Delta Cognitive function=\beta_{0}+\beta_{1}{ADHD-PRS}_{i}+\beta_{2}{Time}_{ij}+\beta_{3}{Sex}_{i}+\beta_{4}{Age}_{i}+\beta_{5}{APOE \varepsilon4}_{i}+\beta_{6}{Years of education}_{i}+\beta_{7}{VRF}_{i}+\beta_{8}{Ancestry}_{i}+\upsilon_{0i}+\varepsilon_{ij}$$

Model 7:

$$\Delta Cognitive function=\beta_{0}+\beta_{1}{ADHD-PRS}_{i} X {Time}_{ij}+\beta_{1}{ADHD-PRS}_{i}+\beta_{2}{Time}_{ij}+\beta_{2}{ADHD-PRS}_{i} X {Sex}_{i}+\beta_{3}{Time}_{ij} X {Sex}_{i}+\beta_{4}{Sex}_{i}+\beta_{5}{ADHD-PRS}_{i} X {Age}_{i}+\beta_{6}{Time}_{ij} X {Age}_{i}+\beta_{7}{Age}_{i}+\beta_{8}{ADHD-PRS}_{i} X {APOE \varepsilon4}_{i}+{\beta_{9}{Time}_{ij} X {APOE \varepsilon4}_{i}+\beta}_{10}{APOE \varepsilon4}_{i}+\beta_{11}{ADHD-PRS}_{i} X {Years of education}_{i}+\beta_{12}{Time}_{ij} X {Years of education}_{i}+\beta_{13}{Years of education}_{i}+\beta_{14}{ADHD-PRS}_{i} X {VRF}_{i}+\beta_{15}{Time}_{ij} X {VRF}_{i}+\beta_{16}{VRF}_{i}+\beta_{17}{Ancestry}_{i}+\upsilon_{0i}+\varepsilon_{ij}$$

Model 8:

$$\Delta Cognitive function=\beta_{0}+\beta_{1}{ADHD-PRS}_{i} X {Time}_{ij}X {A\beta-PET}_{i}+\beta_{2}{ADHD-PRS}_{i}+\beta_{3}{Time}_{ij}+{\beta_{4}A\beta-PET}_{i}+\beta_{5}{ADHD-PRS}_{i} X {Sex}_{i}+\beta_{6}{Time}_{ij} X {Sex}_{i}+\beta_{7}{A\beta-PET}_{i} X {Sex}_{i}+\beta_{8}{Sex}_{i}+\beta_{9}{ADHD-PRS}_{i} X {Age}_{i}+\beta_{10}{Time}_{ij} X {Age}_{i}+{\beta_{11}{A\beta-PET}_{i} X {Age}_{i}+\beta}_{7}{Age}_{i}+\beta_{12}{ADHD-PRS}_{i} X {APOE \varepsilon4}_{i}+{\beta_{13}{Time}_{ij} X {APOE \varepsilon4}_{i}+\beta_{14}{A\beta-PET}_{i} X {APOE \varepsilon4}_{i}+\beta}_{15}{APOE \varepsilon4}_{i}+\beta_{16}{ADHD-PRS}_{i} X {Years of education}_{i}+\beta_{17}{Time}_{ij} X {Years of education}_{i}+\beta_{18}{A\beta-PET}_{i} X {Years of education}_{i}+\beta_{19}{Years of education}_{i}+\beta_{20}{ADHD-PRS}_{i} X {VRF}_{i}+\beta_{21}{Time}_{ij} X {VRF}_{i}+{\beta_{22}{A\beta-PET}_{i} X {VRF}_{i}+\beta_{23}{VRF}_{i}+\beta}_{24}{Ancestry}_{i}+\upsilon_{0i}+\varepsilon_{ij}$$

Model 9:

$$\Delta Cognitive function=\beta_{0}+\beta_{1}{ADHD-PRS}_{i}+\beta_{2}{Time}_{ij}+\beta_{3}{Sex}_{i}+\beta_{4}{Age}_{i}+\beta_{5}{APOE \varepsilon4}_{i}+\beta_{6}{Years of education}_{i}+\beta_{7}{GDS}_{i}+\beta_{8}{Ancestry}_{i}+\upsilon_{0i}+\varepsilon_{ij}$$

Model 10:

$$\Delta Cognitive function=\beta_{0}+\beta_{1}{ADHD-PRS}_{i} X {Time}_{ij}+\beta_{1}{ADHD-PRS}_{i}+\beta_{2}{Time}_{ij}+\beta_{2}{ADHD-PRS}_{i} X {Sex}_{i}+\beta_{3}{Time}_{ij} X {Sex}_{i}+\beta_{4}{Sex}_{i}+\beta_{5}{ADHD-PRS}_{i} X {Age}_{i}+\beta_{6}{Time}_{ij} X {Age}_{i}+\beta_{7}{Age}_{i}+\beta_{8}{ADHD-PRS}_{i} X {APOE \varepsilon4}_{i}+{\beta_{9}{Time}_{ij} X {APOE \varepsilon4}_{i}+\beta}_{10}{APOE \varepsilon4}_{i}+\beta_{11}{ADHD-PRS}_{i} X {Years of education}_{i}+\beta_{12}{Time}_{ij} X {Years of education}_{i}+\beta_{13}{Years of education}_{i}+\beta_{14}{{ADHD-PRS}_{i}X GDS}_{i}+\beta_{15}{{Time}_{ij}X GDS}_{i}+\beta_{16}{GDS}_{i}+\beta_{17}{Ancestry}_{i}+\upsilon_{0i}+\varepsilon_{ij}$$

Model 11:

$$\Delta Cognitive function=\beta_{0}+\beta_{1}{ADHD-PRS}_{i} X {Time}_{ij}X {A\beta-PET}_{i}+\beta_{2}{ADHD-PRS}_{i}+\beta_{3}{Time}_{ij}+{\beta_{4}A\beta-PET}_{i}+\beta_{5}{ADHD-PRS}_{i} X {Sex}_{i}+\beta_{6}{Time}_{ij} X {Sex}_{i}+\beta_{7}{A\beta-PET}_{i} X {Sex}_{i}+\beta_{8}{Sex}_{i}+\beta_{9}{ADHD-PRS}_{i} X {Age}_{i}+\beta_{10}{Time}_{ij} X {Age}_{i}+{\beta_{11}{A\beta-PET}_{i} X {Age}_{i}+\beta}_{7}{Age}_{i}+\beta_{12}{ADHD-PRS}_{i} X {APOE \varepsilon4}_{i}+{\beta_{13}{Time}_{ij} X {APOE \varepsilon4}_{i}+\beta_{14}{A\beta-PET}_{i} X {APOE \varepsilon4}_{i}+\beta}_{15}{APOE \varepsilon4}_{i}+\beta_{16}{ADHD-PRS}_{i} X {Years of education}_{i}+\beta_{17}{Time}_{ij} X {Years of education}_{i}+\beta_{18}{A\beta-PET}_{i} X {Years of education}_{i}+\beta_{19}{Years of education}_{i}+\beta_{20}{{ADHD-PRS}_{i}X GDS}_{i}+\beta_{21}{{Time}_{ij}X GDS}_{i}+\beta_{22}{A\beta-PET}_{i} X {GDS}_{i}+\beta_{23}{GDS}_{i}+\beta_{24}{Ancestry}_{i}+\upsilon_{0i}+\varepsilon_{ij}$$

### **Supplementary Tables and Figures**

#### **Table S1.** **Association between ADHD-PRS and baseline characteristics of the population**

| Continuous variables | β (95% CI) | p-value |
| --- | --- | --- |
| Age, y | -.18 (-.97, .59) | .63 |
| Years of education | -.34 (-.67, -.01) | .04 |
| Years of follow-up | -.01 (-.22, .18) | .85 |
| GDS ^a^ | .03 (-.11, .18) | .67 |
| Categorical variables | OR (95% CI) | p-value |
| Sex ^b^ | .97 (.74, 1.26) | .82 |
| *APOE* ε4 carrier status ^c^ | .80 (.59, 1.08) | .15 |
| VRF burden ^d^ | .85 (.65, 1.12) | .27 |

The association between ADHD-PRS and baseline demographic characteristics was explored using linear or logistic regressions for continuous (age, years of education, years of follow-up, and GDS) or categorical (sex, *APOE* ε4 carrier status, and VRF burden) variables, respectively. All models were corrected to account for any ancestry differences in genetic structure that could bias the results. For that, we used the first seven PCs, as previously recommended for ADNI datasets (2). Abbreviations: y, years; ADHD-PRS, Attention-Deficit/Hyperactivity Disorder polygenic risk score; β, regression coefficient from the linear regression models; CI, confidence interval; *APOE* ε4, apolipoprotein E ε4; VRF, vascular risk factor; GDS, Geriatric Depression Scale; PCs, principal components.

^a^ Depression symptoms were assessed using the GDS scores at baseline.

^b^ Female sex as the reference category.

^c^ Non-carriers as the reference category.

^d^ Categorical variable, high VRF burden was defined as a vascular composite score equal to or higher than two. Decreased VRF burden was used as the reference category.

#### **Table S2. Sensitivity analyses using multiple ADHD-PRS thresholds**

| ADHD-PRS | Model 1:  ADHD-PRS only | | Model 2: Interaction ADHD-PRS with time | | | | | Model 3: Interaction ADHD-PRS with time and Aβ | | | | |
| --- | --- | --- | --- | --- | --- | --- | --- | --- | --- | --- | --- | --- |
| P-value threshold | β (95% CI) | p-value | | | β (95% CI) | p-value | | | | β (95% CI) | p-value | |
| Preclinical Alzheimer Cognitive composite (PACC) | | | | | | | | | | | | |
| ADHD-PRS 0.5 | -.12 (-.44, .19) | .45 | | -.10 (-.17, -.03) | | | .002 | | -.18 (-.32, -.04) | | | .01 |
| ADHD-PRS 0.4 | -.09 (-.41, .22) | .55 | | -.09 (-.16, -.03) | | | .003 | | -.18 (-.33, -.03) | | | .01 |
| ADHD-PRS 0.3 | -.10 (-.43, .21) | .51 | | -.09 (-.16, -.03) | | | .004 | | -.17 (-.32, -.02) | | | .02 |
| ADHD-PRS 0.2 | -.12 (-.44, .20) | .46 | | -.10 (-.17, -.03) | | | .002 | | -.20 (-.34, -.05) | | | .007 |
| ADHD-PRS 0.1 | -.19 (-.52, .12) | .23 | | -.10 (-.17, -.04) | | | .002 | | -.19 (-.34, -.04) | | | .009 |
| ADHD-PRS 0.05 | -.23 (-.58, .10) | .17 | | -.08 (-.15, -.009) | | | .02 | | -.12 (-.28, .02) | | | .11 |
| ADHD-PRS 0.005 | -.11 (-.47, .23) | .51 | | -.03 (-.11, .04) | | | .37 | | .05 (-.10, .20) | | | .51 |
| ADHD-PRS 0.0005 | -.13 (-.49, .21) | .44 | | -.04 (-.12, .03) | | | .27 | | -.14 (-.31, .02) | | | .10 |
| ADNI-Executive function | | | | | | | | | | | | |
| ADHD-PRS 0.5 | -.09 (-.18, -.007) | .03 | | -.003 (-.01, .01) | | | .62 | | -.001 (-.03, .03) | | | .91 |
| ADHD-PRS 0.4 | -.08 (-.17, .0004) | .051 | | -.003 (-.01, .01) | | | .62 | | -.002 (-.03, .03) | | | .88 |
| ADHD-PRS 0.3 | -.08 (-.17, .003) | .06 | | -.003 (-.01, .01) | | | .66 | | .0003 (-.03, .03) | | | .98 |
| ADHD-PRS 0.2 | -.07 (-.16, .01) | .10 | | -.008 (-.02, .007) | | | .28 | | -.01 (-.04, .02) | | | .56 |
| ADHD-PRS 0.1 | -.09 (-.18, -.004) | .03 | | -.01 (-.02, .002) | | | .09 | | -.01 (-.04, .02) | | | .50 |
| ADHD-PRS 0.05 | -.09 (-.18, .001) | .053 | | -.01 (-.03, .003) | | | .12 | | -.02 (-.05, .01) | | | .23 |
| ADHD-PRS 0.005 | -.08 (-.18, .008) | .07 | | -.002 (-.01, .01) | | | .82 | | .02 (-.007, .06) | | | .12 |
| ADHD-PRS 0.0005 | -.04 (-.13, .05) | .38 | | .0001 (-.01, .01) | | | .98 | | .03 (-.001, .07) | | | .06 |
| ADNI-Memory | | | | | | | | | | | | |
| ADHD-PRS 0.5 | .01 (-.05, .08) | .66 | | -.01 (-.02, -.001) | | | .02 | | -.02 (-.05, .0009) | | | .059 |
| ADHD-PRS 0.4 | .01 (-.04, .08) | .60 | | -.01 (-.02, -.002) | | | .01 | | -.02 (-.05, .0007) | | | .057 |
| ADHD-PRS 0.3 | .01 (-.05, .08) | .66 | | -.01 (-.02, -.002) | | | .01 | | -.02 (-.05, -.00006) | | | .04 |
| ADHD-PRS 0.2 | .01 (-.05, .07) | .71 | | -.01 (-.02, -.003) | | | .01 | | -.03 (-.05, -.006) | | | .01 |
| ADHD-PRS 0.1 | .01 (-.05, .08) | .62 | | -.01 (-.02, -.004) | | | .009 | | -.03 (-.06, -.01) | | | .005 |
| ADHD-PRS 0.05 | .01 (-.05, .09) | .58 | | -.01 (-.02, .002) | | | .12 | | -.02 (-.05, .005) | | | .11 |
| ADHD-PRS 0.005 | .06 (-.008, .13) | .08 | | .001 (-.01, .01) | | | .80 | | .01 (-.01, .03) | | | .40 |
| ADHD-PRS 0.0005 | .02 (-.04, .09) | .51 | | .009 (-.004, .02) | | | .16 | | -.02 (-.05, .08) | | | .15 |

Our main analyses were performed with the ADHD-PRS threshold of 1, assuming all genetic markers contributed to ADHD diagnosis. Table S2 shows the results of sensitivity analyses using additional p-value thresholds from 0.5 to 0.0005. The table shows that the main findings were replicated using most ADHD-PRS thresholds, supporting the robustness of our results. All models were adjusted for sex, age at baseline, *APOE* ε4 carriership status (carriers vs. non-carriers), years of education, and ancestry (using the first seven PCs to account for ancestry differences in genetics structure that could bias the results). Abbreviations: ADHD-PRS, Attention-Deficit/Hyperactivity Disorder polygenic risk score; β, regression coefficient from the mixed-effects linear regression models; CI, confidence interval; PACC, Preclinical Alzheimer’s Cognitive Composite; Aβ, amyloid-β; PCs, principal components.

#### **Table S3. Sensitivity analyses controlling for vascular risk factors**

|  | Model 6: ADHD-PRS only | | Model 7: Interaction ADHD-PRS with time | | | | Model 8: Interaction ADHD-PRS with time and Aβ | |
| --- | --- | --- | --- | --- | --- | --- | --- | --- |
|  | β (95% CI) | p-value | | β (95% CI) | p-value | β (95% CI) | | p-value |
| PACC | -.13 (-.46, .18) | .39 | | -.10 (-.17, -.03) | .003 | -.17 (-.31, -.02) | | .02 |
| ADNI-EF | -.10 (-.18, -.01) | .02 | | -.004 (-.02, .01) | .60 | .002 (-.03, .03) | | .88 |
| ADNI-Mem | .01 (-.05, .08) | .65 | | -.12 (-.26, .002) | .055 | -.02 (-.05, -.002) | | .03 |

Table S3 shows that our main findings were replicated adjusting for VRFs. VRF burden was assessed using a composite score. An elevated VRF burden was defined as a vascular composite score equal to or higher than two. All models were also adjusted for sex, age at baseline, *APOE* ε4 carriership status (carriers vs. non-carriers), years of education, and ancestry (using the first seven PCs to account for ancestry differences in genetics structure that could bias the results). Abbreviations: ADHD-PRS, Attention-Deficit/Hyperactivity Disorder polygenic risk score; β, regression coefficient from the mixed-effects linear regression models; CI, confidence interval; PACC, Preclinical Alzheimer’s Cognitive Composite; ADNI-EF, Alzheimer’s Disease Neuroimaging Initiative composite score for executive function; ADNI-Mem, Alzheimer’s Disease Neuroimaging Initiative composite score for memory function; Aβ, amyloid-β; PCs, principal components; VRFs, vascular risk factors.

#### **Table S4. Sensitivity analyses controlling for depression symptoms**

|  | Model 9: ADHD-PRS only | | Model 10: Interaction ADHD-PRS with time | | | | Model 11: Interaction ADHD-PRS with time and Aβ | |
| --- | --- | --- | --- | --- | --- | --- | --- | --- |
|  | β (95% CI) | p-value | | β (95% CI) | p-value | β (95% CI) | | p-value |
| PACC | -.11 (-.43, .20) | .47 | | -.10 (-.16, -.03) | .003 | -.18 (-.32, -.03) | | .01 |
| ADNI-EF | -.09 (-.17, -.005) | .03 | | -.003 (-.01, .01) | .65 | .0008 (-.03, .03) | | .96 |
| ADNI-Mem | .01 (-.04, .08) | .61 | | -.01 (-.02, -.002) | .01 | -.02 (-.05, -.001) | | .03 |

Table S4 shows that our main findings were replicated adjusting for depression symptoms. Depression symptoms were assessed using the GDS scores at baseline. All models were also adjusted for sex, age at baseline, *APOE* ε4 carriership status (carriers vs. non-carriers), years of education, and ancestry (using the first seven PCs to account for ancestry differences in genetics structure that could bias the results). Abbreviations: ADHD-PRS, Attention-Deficit/Hyperactivity Disorder polygenic risk score; β, regression coefficient from the mixed-effects linear regression models; CI, confidence interval; PACC, Preclinical Alzheimer’s Cognitive Composite; ADNI-EF, Alzheimer’s Disease Neuroimaging Initiative composite score for executive function; ADNI-Mem, Alzheimer’s Disease Neuroimaging Initiative composite score for memory function; Aβ, amyloid-β; PCs, principal components; GDS, Geriatric Depression Scale.

#### **Figure S1. Study flow-chart**

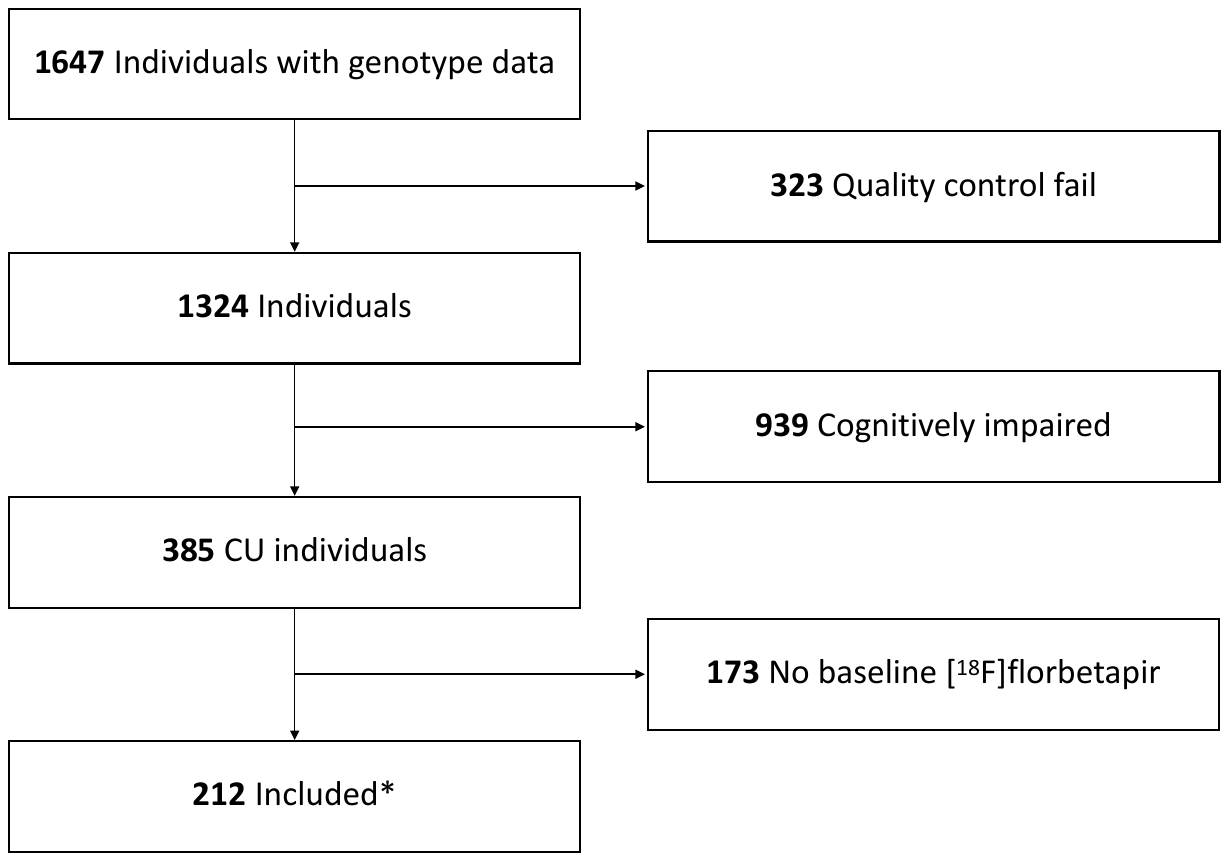

* 16 individuals had no longitudinal MRI data. 18 individuals had no CSF p-tau_181_ data, and one had CSF p-tau_181_ concentrations three standard deviations above the mean.

Abbreviations: CU, cognitively unimpaired; MRI, magnetic resonance imaging; CSF, cerebrospinal fluid.

#### **Figure S2. ADHD-PRS has a normal distribution in the studied population**

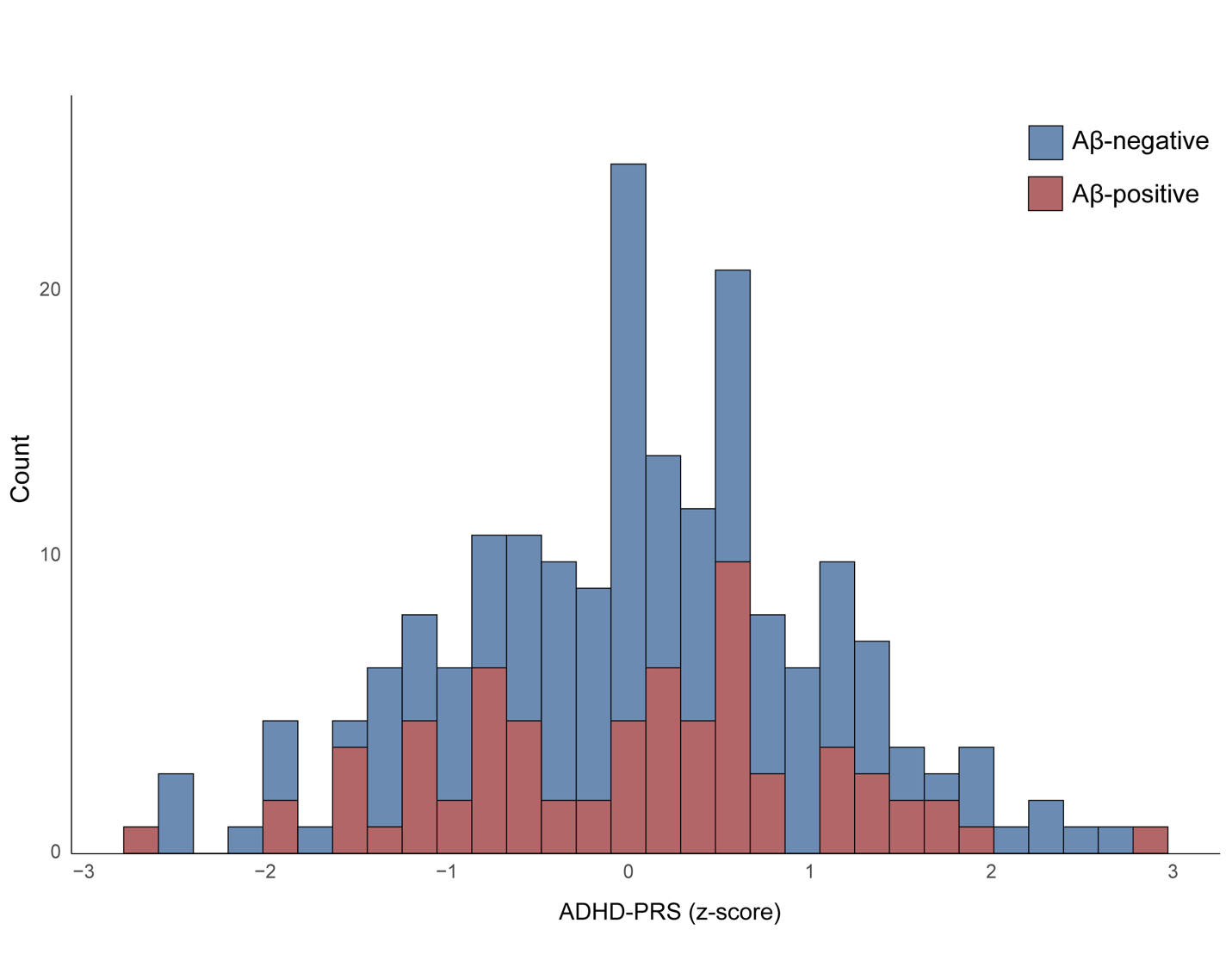

Abbreviations: ADHD-PRS, Attention-Deficit/Hyperactivity Disorder polygenic risk score; Aβ, amyloid-β.

### **References**

1.	Ferrari-Souza JP, Brum WS, Hauschild LA, Da Ros LU, Lukasewicz Ferreira PC, Bellaver B, et al. Vascular risk burden is a key player in the early progression of Alzheimer’s disease. medRxiv [Internet]. 2021 Jan 1;2021.12.18.21267994. Available from: http://medrxiv.org/content/early/2021/12/19/2021.12.18.21267994.abstract

2.	Zettergren A, Lord J, Ashton NJ, Benedet AL, Karikari TK, Lantero Rodriguez J, et al. Association between polygenic risk score of Alzheimer’s disease and plasma phosphorylated tau in individuals from the Alzheimer’s Disease Neuroimaging Initiative. Alzheimers Res Ther [Internet]. 2021;13(1):17. Available from: https://doi.org/10.1186/s13195-020-00754-8
